# Stronger Microstructural Damage Revealed in Multiple Sclerosis Lesions with Central Vein Sign by Quantitative Gradient Echo MRI

**DOI:** 10.1101/2021.07.17.21260663

**Authors:** Victoria A. Levasseur, Biao Xiang, Amber Salter, Dmitriy A. Yablonskiy, Anne H. Cross

## Abstract

**Background:** Multiple sclerosis (MS) lesions typically form around a central vein that can be visualized with FLAIR* MRI, creating the central vein sign (CVS) which may reflect lesion pathophysiology. Herein we used Gradient Echo Plural Contrast Imaging (GEPCI) MRI to simultaneously visualize CVS and measure tissue damage in MS lesions. We examined CVS in relation to tissue integrity in white matter (WM) lesions and among MS subtypes.

**Subjects and Methods:** Thirty relapsing–remitting MS (RRMS) subjects and 38 progressive MS (PMS) subjects were scanned with GEPCI protocol. GEPCI T2*-SWI images were generated to visualize CVS. Two investigators independently evaluated WM lesions for CVS and measured lesion volumes. To estimate tissue damage severity, total lesion volume, mean lesion volume, R2t*-based tissue damage score (TDS) of individual lesions and tissue damage load (TDL) were measured for CVS+, CVS-, and confluent lesions. Spearman correlations were made between MRI and clinical data. A paired t-test was used to compare measurements of CVS+ versus CVS- lesions in each individual.

**Results:** 398 of 548 lesions meeting inclusion criteria showed CVS. Most patients had ≥ 40% CVS+ lesions. CVS+ lesions were present in similar proportion among MS subtypes. Interobserver agreement was high for CVS detection. CVS+ and confluent lesions had higher average and total volumes versus CVS- lesions. CVS+ and confluent lesions had more tissue damage than CVS- lesions based on TDL and mean TDS.

**Conclusion:** CVS occurred in RRMS and PMS in similar proportions. CVS+ lesions had greater tissue damage and larger size than CVS- lesions.

## Introduction

Multiple sclerosis (MS) is a common inflammatory neurologic disease that affects at least 620,000 individuals in the United States alone.^1^ Worldwide estimates of disease prevalence approach 2.5 million.^2^ Non-invasive imaging techniques are useful tools to help elucidate underlying pathology of diseases of the central nervous system (CNS). Magnetic resonance imaging (MRI) is integral to MS diagnosis and has provided valuable insights into disease pathology and evolution. Advancements in quantitative MRI imaging have elucidated a more detailed picture of MS lesions compared to standard clinical imaging. Furthermore, the development of novel imaging techniques and better measurements of tissue integrity may help to improve our understanding of the relationship between clinical disability and lesion burden,^3^ as well as distinguish MS from other diseases.

MS lesions, especially those in the white matter (WM), typically form around a central vein.^4^ The perivenular area contains antigen presenting cells that can activate cells of the adaptive immune system, with subsequent lesion formation.^5^ This lesion topography creates the central vein sign (CVS) on MRI.^6^ Ultra high field MRI and post-processing techniques that capture the relationship between venous structures and T2-weighted hyperintensities can help to differentiate MS from diseases with similar appearing lesions.^7^ It has been proposed that having more than 40% lesions with CVS or three or more lesions with CVS is specific for MS.^8^ Other studies have proposed varying CVS thresholds and absolute lesion number, with similar degrees of specificity.^7,9^ Advanced post-processing techniques, such as T2* and FLAIR* improve discernment of CVS.^10,11,12^

In this study we used a gradient echo plural contrast imaging (GEPCI) technique that is based on a multi-echo gradient echo MR sequence and post processing algorithms, to generate images and quantitative maps with different biological contrasts.^13,14^ T2*-SWI images, which combine GEPCI susceptibility weighted imaging (SWI) with T2* (T2* = 1/R2*) maps were used to visualize venous structures within T2* hyperintensities. Because images with different contrasts obtained using GEPCI are naturally co-registered, T2*-SWI images are especially useful for localizing central veins within MS lesions. Making use of recently-developed postprocessing algorithms,^15,16^ we also generated quantitative tissue-specific R2t* maps (t represents “tissue cellular” specific component of R2*) allowing the estimatation of CNS tissue damage due to MS.^18,19,20^ The tissue cellular specific component R2t* was separated from total R2* by removing the contribution of blood oxygen level-dependent effects.^15^

We examined relationships of lesions with and without CVS to the underlying tissue damage based on R2t*, and to MS subtype and disability measurements. We hypothesized that lesions with CVS would reflect regions with enhanced underlying adaptive immune responses and greater tissue damage than those without CVS.

## Materials and Methods

### Study design

This was a single-center cross-sectional study to compare the presence of CVS in people with RRMS and non-relapsing PMS, to assess tissue damage in lesions with and without CVS using R2t* as an estimate of underlying tissue integrity, and to determine correlations of CVS burden with clinical disability.

### Subjects

Sixty-eight subjects (30 relapsing and 38 progressive) ≥ 18 years of age diagnosed with relapsing-remitting, primary progressive, or secondary progressive MS and able to provide written informed consent were included. Clinical subtypes were assigned by clinicians prior to the study and in advance of imaging, based on neurological assessement and disease course. RRMS and PMS subjects were recruited to be of similar ages, to remove the confounder of age when comparing clinical subtypes. PMS subjects were required to be progressing in absence of inflammatory disease activity. RRMS subjects had no progression unless directly related to MS attacks. The presence of hypertension and hyperlipidemia was also noted.

### MRI protocol and post-processing

Magnetic resonance data were acquired on a 3T Trio MRI scanner (Siemens, Erlangen, Germany) using a 32-channel phased-array head RF coil. GEPCI data were acquired using three-dimensional multi-gradient-echo sequence with flip angle 30°, TR=50ms, voxel size 1×1×2mm^3^ and acquisition time 12 minutes. Ten gradient echoes, with first echo time TE1=4ms and echo spacing ΔTE=4ms were collected. Effects of physiological fluctuations were also mitigated using previously developed technique.^21^ Fluid-attenuated inversion recovery (FLAIR) images (voxel size 1×1×3mm^3^) were also acquired. Procedures of image reconstruction and generating T2*-SWI and R2t* images were described previously.^13,15^ Both R2* and R2t* maps were generated accounting for adverse effects of magnetic field inhomogeneities using voxel spread function approach.^22^ T2*-SWI images were reconstructed into 0.5×0.5×1mm^3^ for visualizing lesions with CVS. Image reconstruction and post-processing of GEPCI data were performed using a standard PC computer and Matlab software (Math-Works Inc.). For each subject, total lesion volume, mean lesion volume, tissue damage load (TDL) and mean tissue damage score (TDS) were measured for lesions with CVS (CVS+), lesions without CVS (CVS-) and confluent lesions. Lesion volumes of CVS+ lesions were measured by summing the volumes of all CVS+ lesions in a subject. Mean lesion volume in CVS+ lesions were calculated as lesion volume of CVS+ lesions divided by number of CVS+ lesions in a subject. Total lesion volume and mean lesion volume of CVS- and confluent lesions were calculated in the same way as CVS+ lesions. Mean TDS of CVS+/-lesions were defined as:

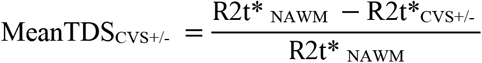

Where R2t* _NAWM_ is the median value of R2t* in normal appearing white matter (NAWM) in that individual and R2t*_CVS+/-_ is the median value of R2t* in CVS+/- lesions. The center voxel, corresponding to the vein, was excluded while computing R2t*_CVS+_. Exclusion of center voxels and using median values minimized the effect of venous structures on R2t*_CVS+_ calculation. Higher mean TDS reflects more severe tissue damage. Tissue Damage Load (TDL) in CVS+/- lesions was defined as:

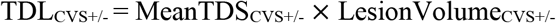

Mean TDS and TDL of confluent lesions were calculated in the same way as CVS+/-lesions. Note that these measures in CVS- lesions are different from CVS+ lesions only by keeping the center voxel. Six patients did not have CVS- lesions; their volume of CVS- lesions was assigned as 0. All patients had lesions characteristic of MS.

### Central vein sign detection

Dual reader evaluation of lesion volume, anatomic location, and presence or absence of CVS were recorded using imaging analysis software ITK-SNAP.^23^ CVS was determined on T2*-SWI images, which inherently co-localize T2*-hyperintensities with venous structures (**Figure 1**). Central vein was identified using criteria previously.^8^ Veins were required to appear as a thin line or dot, detectable in at least two perpendicular planes, appearing as a thin line in one plane and located in the middle of the WM lesion. The diameter of the vein was less than 2mm, and crossed partly or completely through the lesions.^8^ The total number and proportion (CVS%) of lesions with CVS were calculated. Inclusion criteria for CVS were cerebral lesions in WM, including the juxtacortical regions and brainstem. Lesions were excluded if they were less than 3 mm diameter in any plane, confluent, included more than 1 vein, or difficult to decipher due to artifact.^8^ Since GEPCI imaging did not cover the entire cerebellum, cerebellar lesions were excluded. Lesion location (cortical/juxtacortical, subcortical, periventricular, brainstem) was documented. 548 WM lesions meeting inclusion criteria and 143 confluent lesions were identified by a neurologist (VAL) and assessed separately by a second experienced reader (BX) to formulate inter-rater reliability. In cases of disagreement (<10% of lesions), a third reader (AHC) adjudicated.

**Figure 1.**
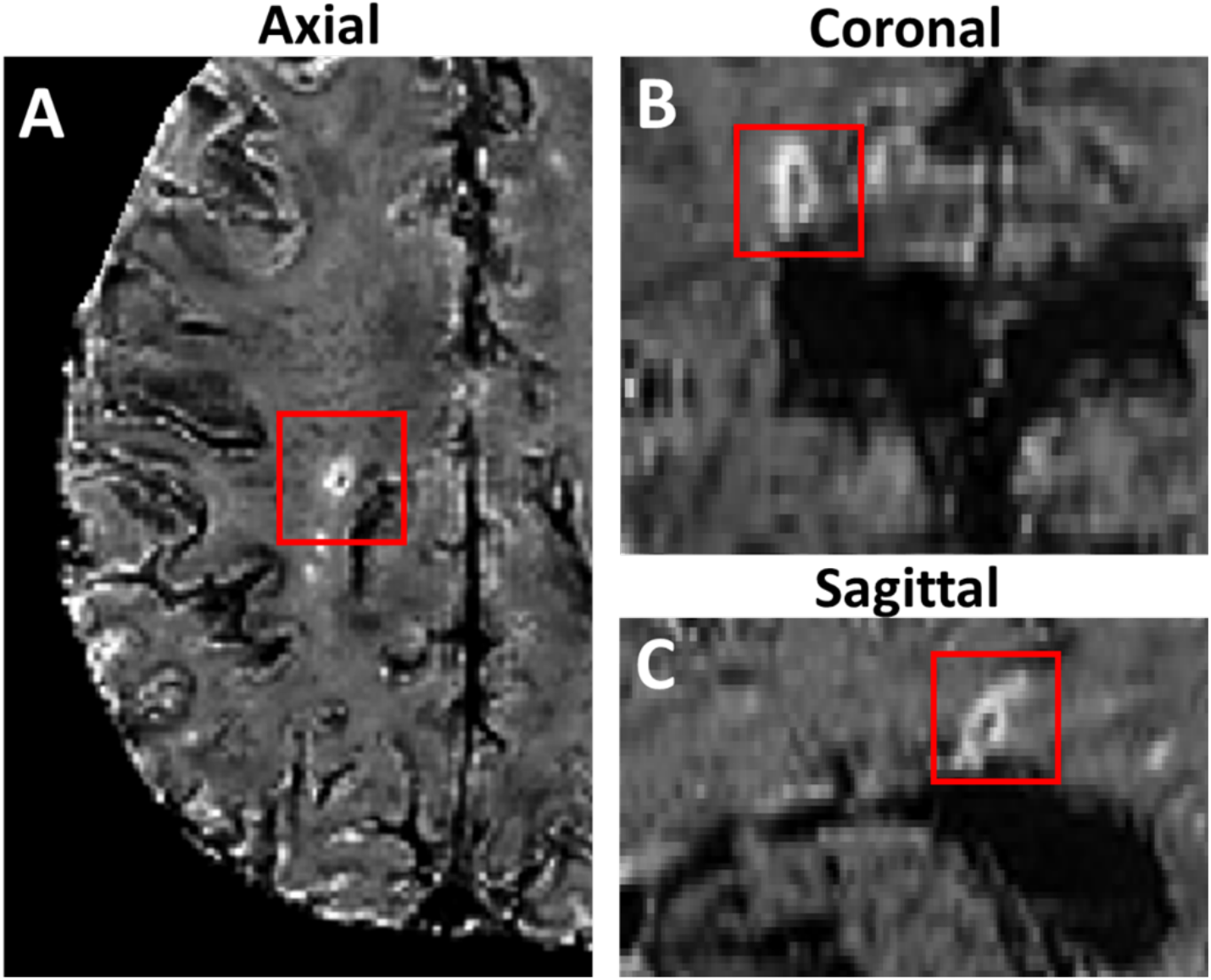
T2*-SWI images (0.5 × 0.5 × 1 mm^3^ resolution) of the brain of a person with RRMS demonstrate co-localization of WM lesion with central vein, highlighted in axial, coronal and sagittal planes (**A-C**).

### Neurological examinations

On the day of imaging, subjects underwent the following tests: 25 foot time walk (25FTW), nine hole peg test (9HPT), 2-second Paced Auditory Serial Addition Test (PASAT-2), 3-second PASAT (PASAT-3), Symbol Digit Modality Test (SDMT, oral version), and Expanded Disability Status Score (EDSS).

### Data analysis

Differences in the proportion of CVS+ lesions between the RRMS and PMS subjects were evaluated using the Wilcoxon sign rank test. Cohen’s kappa statistic was used to measure interrater reliability (κ<0, poor agreement; 0<κ<0.2, slight agreement; 0.2<κ<0.4, fair agreement; 0.4<κ<0.6, moderate agreement; 0.6<κ<0.8, substantial agreement; and 0.8<κ<1, almost perfect agreement).^24^ Intraclass correlation coefficient (ICC) was used for lesion volume measurement reliability. Lesion volume, TDL, and TDS were compared between CVS+, CVS- and confluent lesions. Paired t-tests were used to compare lesion volume, mean lesion volume, TDL, and mean TDS measurements of CVS+ and CVS- lesions. False discovery rate was used to adjust for multiple comparison. Spearman’s rank-order correlation was used to investigate the correlation between CVS proportion and clinical scores.

## Results

### Patient Characteristics, Lesion Counts, and Lesion Distribution

Clinical characteristics of the 30 RRMS and 38 PMS subjects are shown in **Table 1**. Subjects in the RRMS group and in the PMS group were specifically enrolled to be of similar age range to prevent confounding differences due to age. PMS participants had significantly higher (worse) EDSS scores and lower SDMT, and MSFC scores compared to the RRMS group (p < 0.001). PASAT (2sec) scores were also lower in the PMS group (p < 0.05). Although nominally worse in the PMS group, differences in PASAT (3sec) scores were not statistically significant between subtypes. 548 WM lesions meeting CVS inclusion criteria and 143 confluent lesions were analyzed in 68 participants. Of the 548 lesions, most of them were identified in the periventricular (n = 162), juxtacortical/cortical (n = 270), and subcortical (n = 98) regions (**Table 2**). Lesions in the periventricular region showed higher percentage of CVS than lesions in juxtacortical/cortical, subcortical and brain stem regions. Our data showed that 91.2% of participants had ≥ 40% CVS+ lesions, a threshold proposed to distinguish MS from other CNS inflammatory disease.^8,25,26^

**Table 1:** Mean group differences between RRMS and PMS subjects. Subjects with RRMS and PMS were recruited to be of similar ages. Participants with PMS did worse on most tests, with significantly higher EDSS scores and lower SDMT, PASAT 2sec, and MSFC scores than RRMS participants. CVS+ lesion total was not different between clinical subtypes. Values Presented as Mean ± SD, p-values: a=ANOVA, b=Chi-square. **\***Missing values: Disease duration = 2, MSFC = 2.

| **Factor** | **Total**<br>**(N=68)** | **RRMS**<br>**(N=30)** | **PMS**<br>**(N=38)** | **p-value** |
| --- | --- | --- | --- | --- |
| **Sex (Male/Female)** | 21/47 | 5/25 | 16/22 | **< 0.001<sup>b</sup>** |
| **Age** | 59.6±9.5 | 57.6±10.3 | 61.2±8.6 | 0.12<sup>a</sup> |
| **Disease Duration\*** | 18.4±9.3 | 18.6±9.1 | 19.1±10.1 | 0.53<sup>a</sup> |
| **EDSS** | 4.2±1.9 | 2.6±1.2 | 5.5±1.4 | **<0.001<sup>a</sup>** |
| **SDMT Oral Score** | 47.6±13.3 | 54.1±9.3 | 42.4±13.8 | **<0.001<sup>a</sup>** |
| **PASAT 2sec** | 31.5±10.4 | 34.4±9.0 | 29.2±11.0 | **0.038<sup>a</sup>** |
| **PASAT 3sec** | 43.2±12.3 | 46.1±10.1 | 41.0±13.4 | 0.085<sup>a</sup> |
| **MSFC\*** | -1.08±2.4 | 0.26±1.03 | -2.2±2.6 | **<0.001<sup>a</sup>** |

**Table 2.**
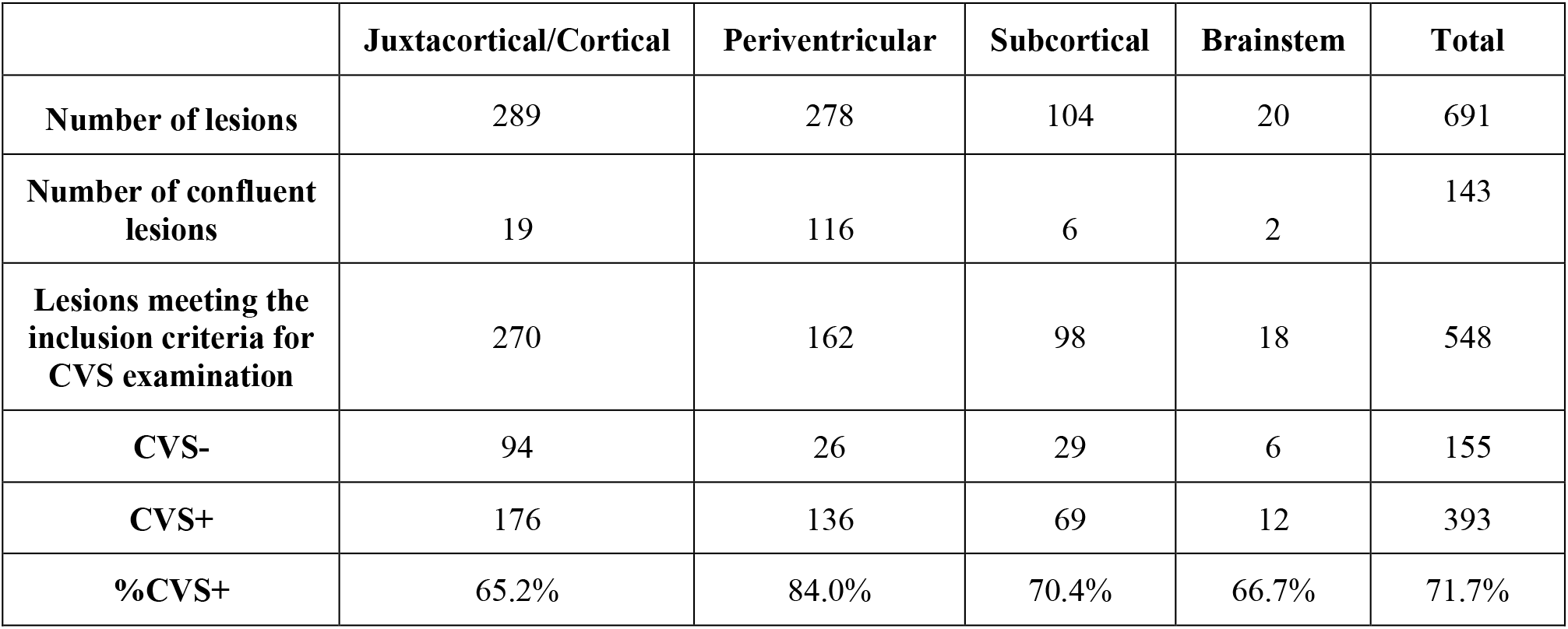
Distribution of lesions examined. Lesion location in relations to presence or absence of CVS, and percentage of lesions with CVS were recorded. Veins were required to appear as a thin line or dot, detectable in at least two perpendicular planes, appearing as a thin line in one plane and located in the middle of the WM lesion. The diameter of the vein was less than 2mm, and crossed partly or completely through the lesions.

CVS was detected on T2*-SWI in axial, coronal and sagittal planes (**Figure 1**). TDS, calculated from R2t*, was used to quantify tissue damage in lesions (**Figure 2**). Tissue damage score was not significantly different amoung lesions in juxtacortical/cortical, subcortical, periventricular or brainstem regions (**supporting information, figure 1S**).

**Figure 2.**
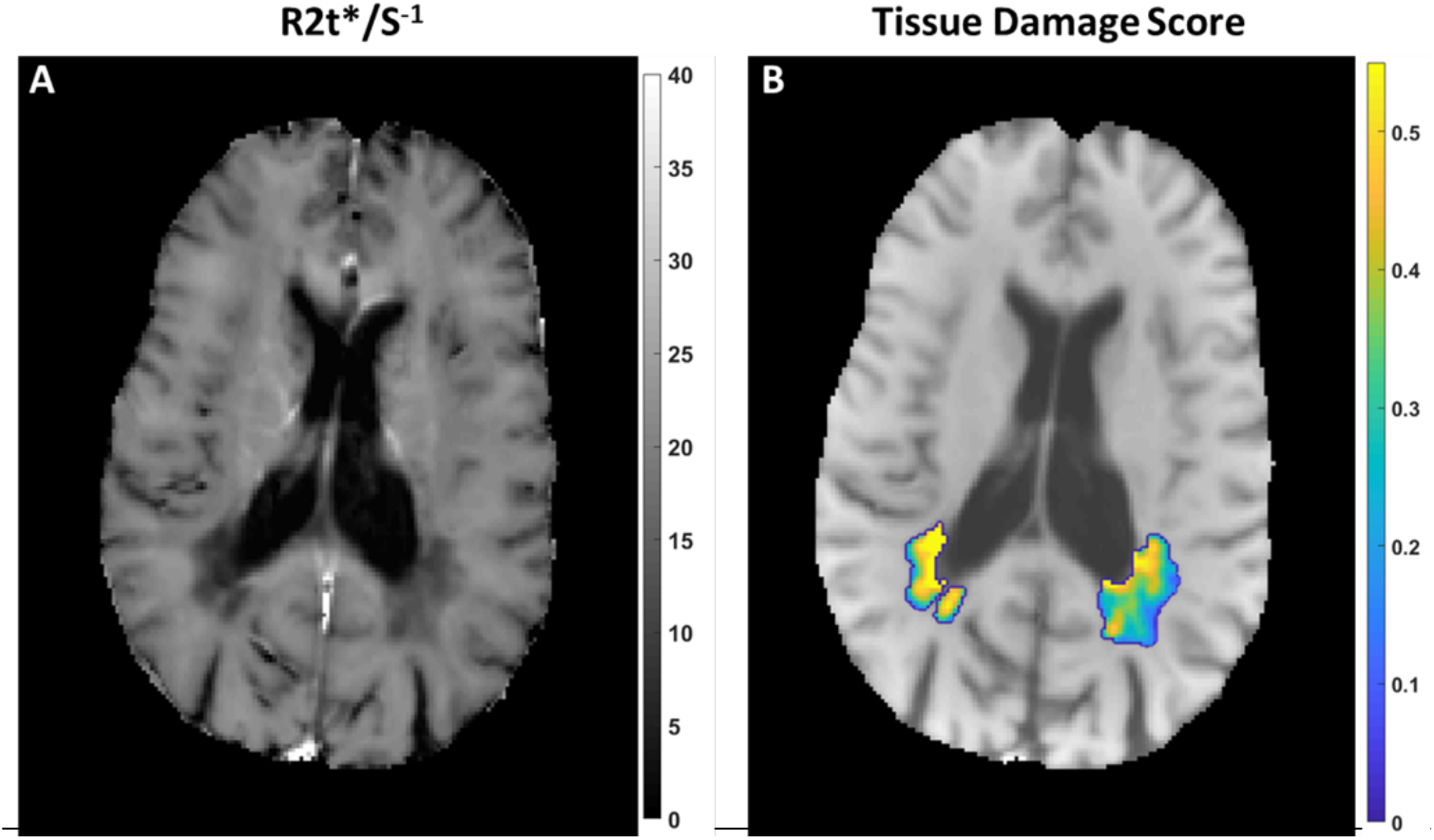
R2t* and tissue damage score (TDS) maps show two periventricular lesions of a person with PMS. Color coded R2t*- based TDS is superimposed on the GEPCI-T1w image. TDS map (B) shows the heterogeneity of damage in the two lesions. Higher TDS represents lower R2t*, indicating more severe tissue damage.

### Interobserver agreement

Two readers independently evaluated 548 WM lesions for CVS analysis, measurement of lesion volumes including volumes of an additional 143 confluent lesions. Interrater reliability of lesion measurement was high (ICC = 0.99, 95% **Figure 3**). Cohen’s kappa for interrater reliability showed substantial agreement for CVS reading between two raters (VAL and BX) (κ=0.72). The two readers had different classifications of CVS+ versus CVS-for 50 out of 548 lesions (9.1%). The third reader (AHC) adjudicated these 50 lesions.

**Figure 3.**
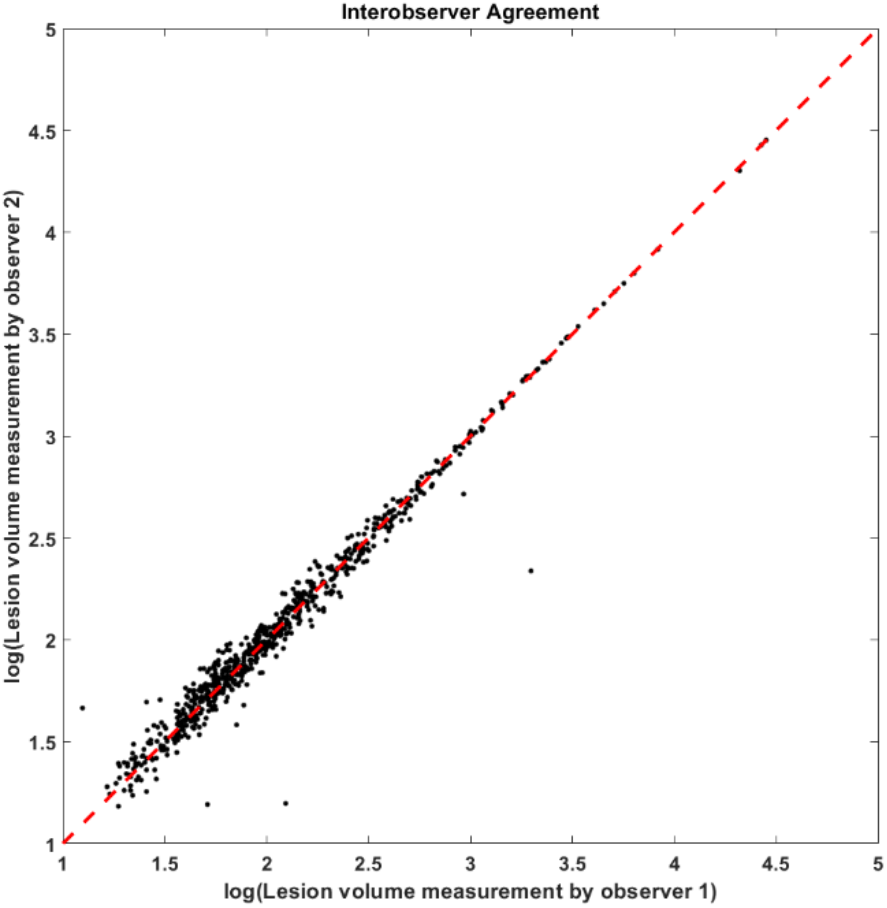
Correlation analysis of independent lesion volume (mm^3^) measures by two investigators, R^2^ = 0.96.

### Lesions with CVS had greater volumes

CVS+ lesions had higher total volumes (**Figure 4**, top row) and mean volumes (**Figure 4**, bottom row) when compared with CVS- lesions. This was true for all subjects when combined (**Figure 4**, left column), and for PMS and RRMS subtypes individually (**Figure 4**). Confluent lesions showed significantly higher total and mean volumes than CVS- and CVS+ lesions (p<0.001).

**Figure 4.**
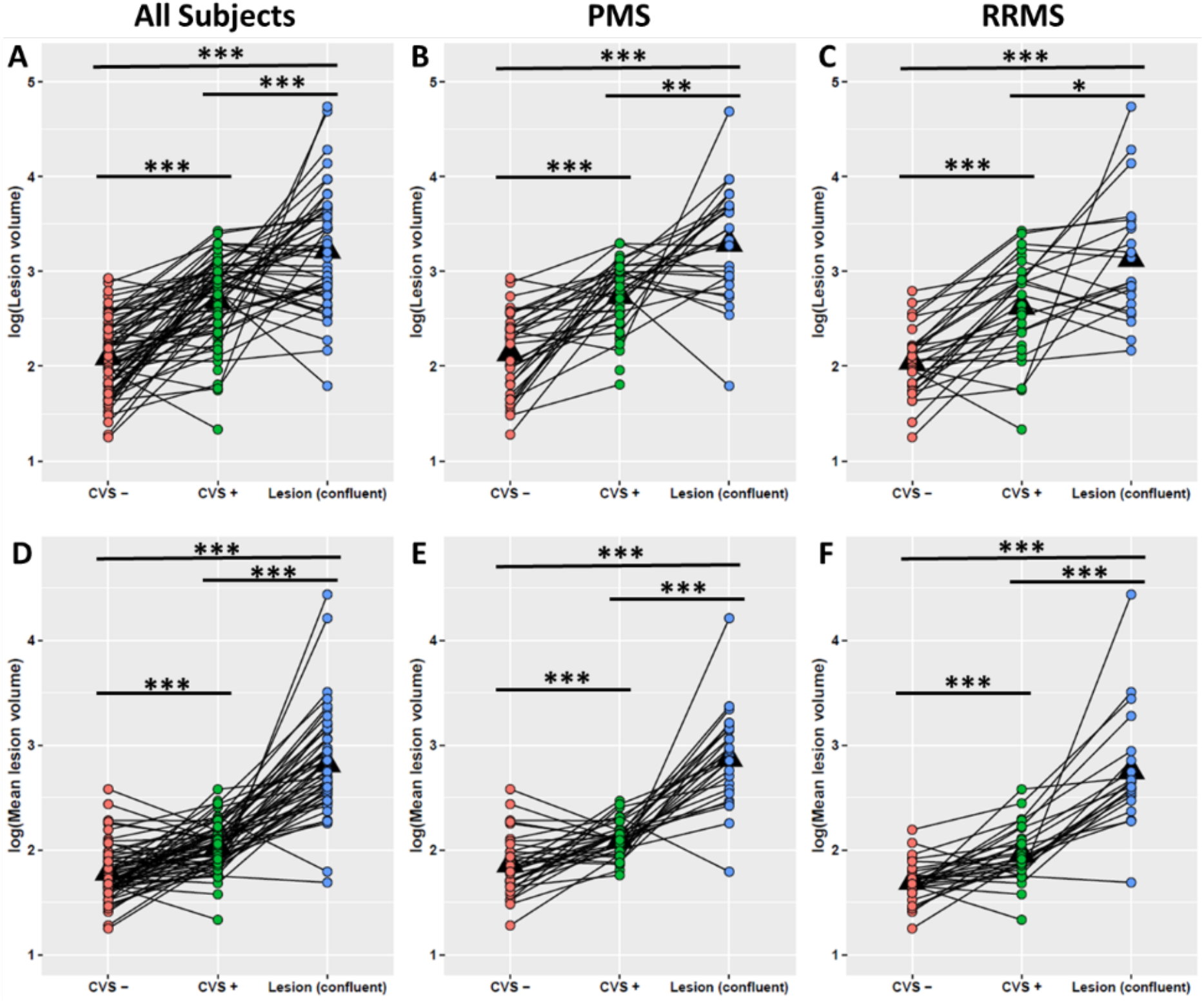
Paired t-tests comparing total lesion volume (A-C) and mean lesion volume (D-F) of lesions with and without CVS and confluent lesions in all subjects (n=68) and separately in PMS and RRMS subgroups. Lesions with CVS (CVS+, green dots) had significantly higher lesion volumes compared to lesions without CVS (CVS-, red dots) in the entire MS group (p < 0.001) and in each subtype (p < 0.001) (**A-C**). Lesions with CVS had significantly higher mean lesion volumes compared to lesions without CVS in the entire MS group (p<0.001) (D), PMS (p<0.001) (E) and RRMS (p<0.001) (F). <u>Note</u>: if a patient did not have CVS- lesions (n = 6), the CVS- lesion volume was assigned as 0. The lines connecting each dot represent data from the same subject. *** p<0.001, ** p<0.01, * p<0.05. All listed p values are after multiple comparison correction using false discovery rate.

### R2t* based damage estimates were greater in CVS+ lesions

R2t* was used to estimate tissue integrity. CVS+ lesions had significantly more tissue damage, as measured by R2t*-based tissue damage load (TDL) and mean tissue damage score (TDS), compared with CVS- lesions (**Figure 5**). CVS+ and confluent lesions showed higher TDL and mean TDS than CVS- lesions (p<0.001). No significant difference in mean TDS was found between CVS+ lesions and confluent lesions.

**Figure 5.**
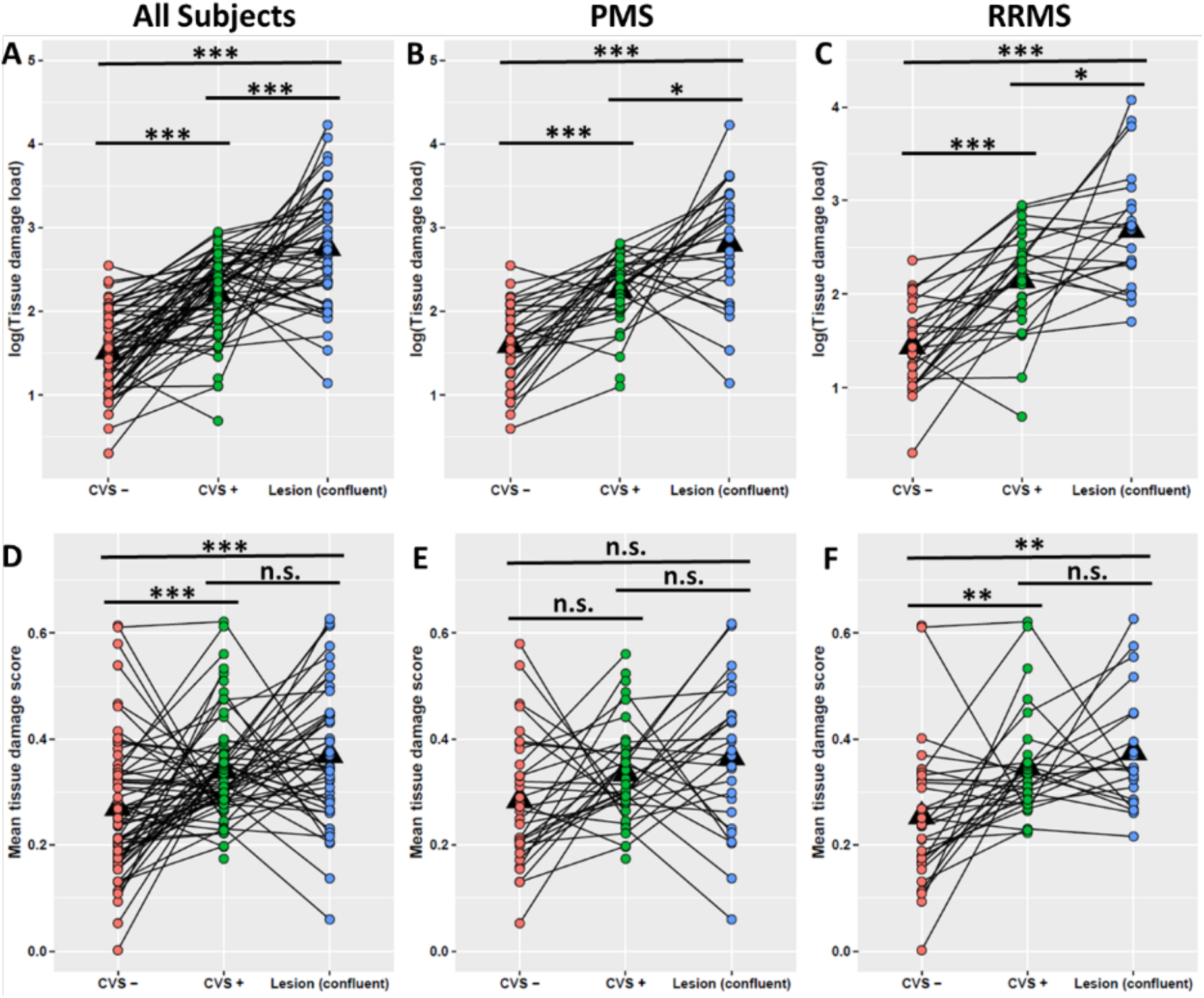
Paired t-tests comparing tissue damage load (TDL) (A-C) and mean tissue damage score (TDS) (D-F) of lesions with and without CVS and confluent lesions in all subjects (n=68), PMS, and RRMS patients. Lesions with CVS (green dots) and confluent lesions (blue dots) had significantly higher TDL and mean TDS compared to lesions without CVS (red dots) in the MS group (p<0.001) (A, D) and the RRMS subtype (p < 0.01) (C, F). <u>Note</u>: if a patient did not have CVS- lesions (n = 6), the CVS- lesion volume was assigned as 0. The lines connecting each dot represent data from the same subject. *** p<0.001, ** p<0.01, * p<0.05, n.s. p≥0.05. All listed p values are after multiple comparison correction using false discovery rate.

### Significant correlations between CVS+ lesion proportion and clinical scores

Correlations between percentage of CVS+ lesions and clinical scores were investigated. After FDR was applied, the percentage of WM lesions with CVS showed significant correlation with EDSS (r=0.37, p=0.002), 25-foot-walk (r=0.29, p=0.018) and 9-hole-peg test (r=0.26, p =0.033).

### No significant difference in lesion counts or proportion of CVS+ lesions between RRMS and PMS subgroups

Average CVS+ counts were not significantly different between subtypes with RRMS having 8.7 and ± 5.2 and PMS with 8.3 ± 4.6 (p = 0.74). The Wilcoxon sign rank test showed no significant difference of CVS% between MS subtypes, although PMS subjects trended toward greater proportion of CVS+ lesions versus RRMS subjects with median CVS% for PMS of 0.80 ± 0.20 and for RRMS of 0.67 ± 0.16 (p=0.077) (**Figure 6**).

**Figure 6.**
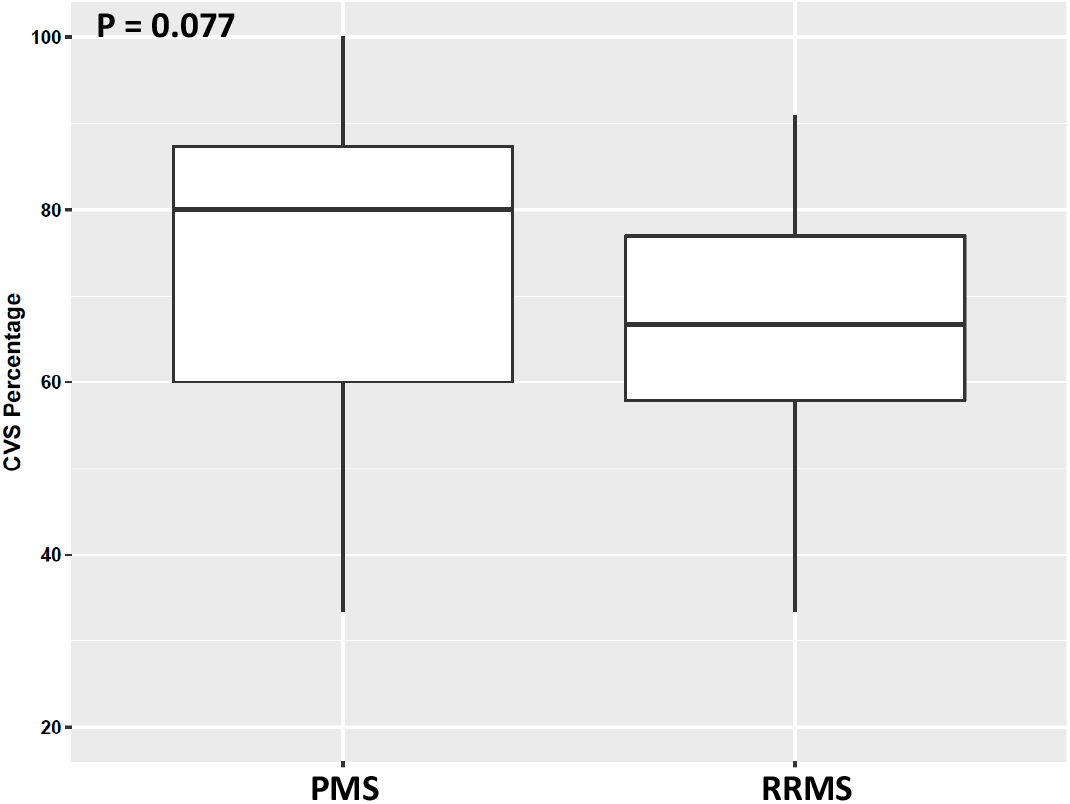
Distribution of proportion of lesions with CVS by MS subtype. There was no significant difference of CVS% between MS subtypes (p=0.077). Boxes represent the interquartile ranges; the horizontal lines within the boxes indicate median values.

### Relationship between hypertension, hyperlipidemia and CVS proportion

White matter hyperintensities are frequently noted on T2w brain images of people with vascular disease. Thus, we compared CVS proportion in subjects with and without hypertension and with and without hyperlipidemia but found no significant differences. Data showed a small but insignificant (p = 0.066) trend toward people with hypertension having larger proportion of lesions with CVS.

## Discussion

In this study, we examined the proportion of lesions with CVS among 68 people with MS, including age-matched relapsing-remitting and progressive MS subtypes. Cohen’s kappa showed substantial inter-rater agreement in this study, similar to previous studies.^11^ The purpose of our study was to examine associations of underlying tissue integrity in lesions with the presence of CVS, identify any differences in the CVS+ lesion proportions between relapsing and progressive MS subtypes and to determine if presence or proportion of lesions with CVS correlated with clinical disability measurements.

High rates of MS misdiagnoses have been reported^27,28^ and thus the use of CVS to improve diagnostic specificity has become increasingly important. Our results demonstrated that T2*-SWI GEPCI-based images provide a reliable method for CVS detection. Importantly, GEPCI allows not only detection of CVS but also the generation of R2t* maps from the same data to provide a quantitative estimate of tissue damage. Out data show that CVS+ lesions detected by T2*-SWI tended to be larger and had worse underlying tissue damage based on R2t* than CVS- lesions.

Herein, tissue damage was measured by the GEPCI quantitative tissue-specific R2t* metric that has been associated with tissue damage pathologically.^19^ Previously, our group also showed that R2t* can detect both gray and white matter alterations in MS.^17,18,19^ The current results imply that lesions with a central vein have worse underlying tissue damage than those lacking a demonstrable central vein, suggesting the possibility of different pathophysiologies between CVS+ and CVS- lesions. The higher TDS in CVS+ than CVS- lesions is unlikely to be due to different prevalence of their topographic distribution (e.g., CVS+ lesions have been demonstrated to be more frequent in periventricular WM), since CVS+ lesions in different regions showed similar TDS **(supporting information, figure 1S**). Acute MS lesions that form around a central vein are characterized by inflammatory cell infiltration, as well as immunoglobulin and complement deposition. Strong associations of axonal loss with degree of cellular infiltration have been reported.^29^ Thus, the presence of a central vein may signify a more destructive disease process than in CVS-MS lesions. MS lesions with CVS may not only be useful for diagnostic accuracy, but also for understanding disease pathophysiology.

It has been hypothesized that CVS- lesions may represent non-demyelinating pathology, including being due to other factors such as vascular disease. A prior study reported that the proportion of CVS+ lesions was significantly decreased in MS patients with hypertension.^30^ Clinical evaluation of patients with non-specific white matter lesions who have comorbid cerebrovascular disease is not straightforward. It is often difficult to distinguish extensive periventricular white matter changes secondary to demyelination from sequelae of vascular disease.^31^ Results from the current study did not show increased CVS- lesion prevalence in those with vascular risk factors.

We found no significant differences in the presence and proportions of lesions with CVS in relapsing versus progressive MS subtypes, in accord with previous studies.^6,32^ These findings are consistent with the CVS reflecting a common lesion pathogenesis among MS subtypes.^32^ Of note, PMS subjects trended toward greater proportion of CVS+ lesions versus RRMS subjects.

Significant correlations between percentage of CVS and clinical disability were found. One previous report showed a relationship between CVS and PASAT (3sec) and California Verbal Learning Test.^25^ This finding supports the present finding of worse underlying tissue damage in CVS+ lesions. Application of CVS status as an imaging biomarker of MS disease severity should be further investigated, including in longitudinal studies.

## Limitations

Although this study was limited by a small sample size (n = 68), the number of lesions studied herein was close to 700. Since patients in the RRMS group were age-matched with individuals with PMS, the MS patients studied were older and with longer disease duration than in most other studies of CVS in MS, which could limit generalizability. We found that this older MS population displayed a similar high proportion of lesions with central veins to younger MS patients reported in the literature.^8^ The present data indicate that the applicability of CVS to aid MS diagnosis is not limited by patient age.

This study may have been underpowered to show increased CVS- lesion in patients with vascular co-morbiditied. Future studies should be performed to further elucidate the relationship of MS lesions with and without CVS to vascular risk factors and to disability measures.

CVS+ lesions were found to be larger than CVS- lesions in this study, raising the possibility that venules within small MS lesions may have been missed due to insufficient image resolution. Image resolution at 3T limited our ability to differentiate cortical lesions, and visualize brainstem structures for analysis. However, even studies at 7T showed that some MS lesions lack CVS.^33^

## Conclusions

Here we have described a technique incorporating a quantitative T2* mapping for CVS detection in brain lesions with simultaneous evaluation of tissue damage in CVS-associated lesions. Importantly, this study also found that CVS+ lesions are associated with more underlying tissue damage and with more MS-specific disability. The present study suggests that lesions without CVS may be fundamentally different than lesions with CVS, and that CVS- lesions contribute less to MS-specific disability. In future studies we will explore the longitudinal relationship between CVS, R2t* and clinical disability, which may help address this question.

## Data Availability

Data used in this manuscript will be available upon reasonable request.

## Ethics Statement

We take full responsibility for the data, the analyses, interpretation, and the conduct of the research. Data used in this manuscript will be available upon reasonable request. We confirm that this research was conducted according to the World Medical Association Declaration of Helinski and conforms to the ICMJE recommendations for the conduct, reporting, editing, and publication of scholarly work in medical journals. The Institutional Review Board at Washington University in St. Louis, Missouri approved the study (IRB ID #: 201412138). A waiver of consent was obtained to access already acquired clinical data (IRB ID #: 202005043; IRB ID #: 202005044).

## Author Contributions

**Victoria A. Levasseur:** Concept and design, leading role in writing of the manuscript, data collection, data analysis/interpretation, leading role in review of literature.

**Biao Xiang:** Concept and design, writing of the manuscript, leading role in data analysis and interpretation, review of literature.

**Amber Salter:** Supporting role in reviewing and editing the manuscript, supporting role in data analysis/interpretation.

**Anne H. Cross:** Study supervision, concept and design, writing of the manuscript, funding acquisition, data interpretation.

**Dmitriy Yablonskiy**: Study supervision, concept and design, writing of the manuscript, funding acquisition, data interpretation.

