## supporting information, figure 1S for "Stronger Microstructural Damage Revealed in Multiple Sclerosis Lesions with Central Vein Sign by Quantitative Gradient Echo MRI"

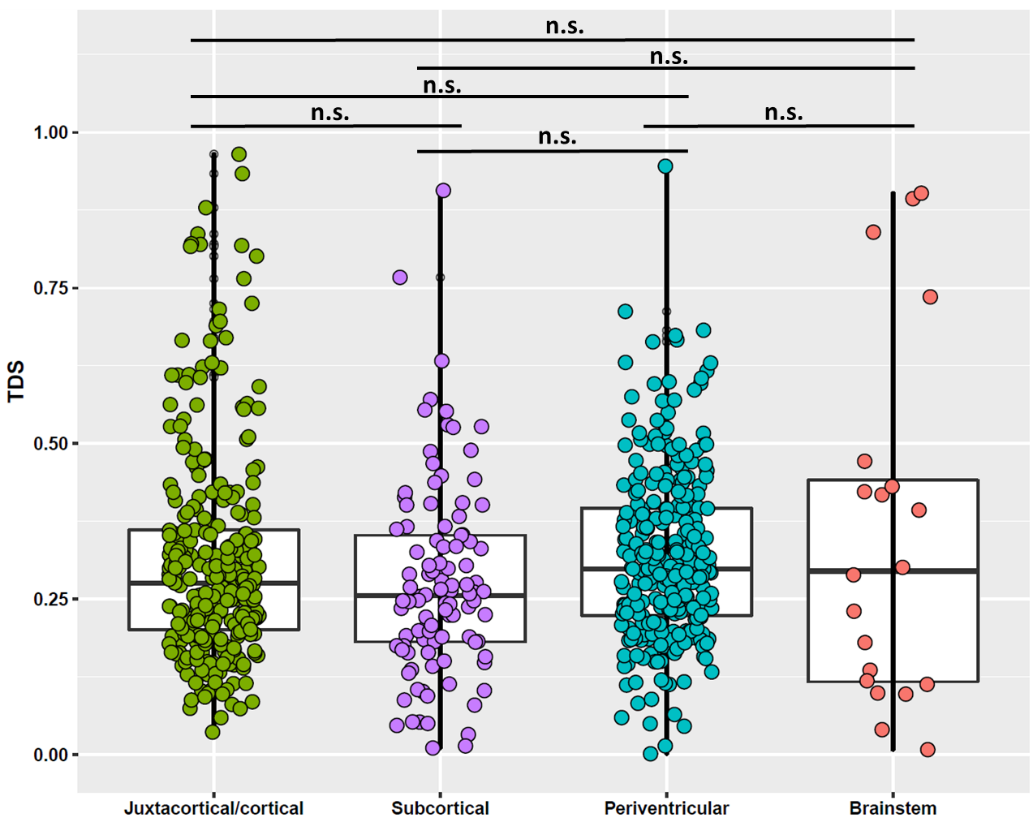

Figure 1S. Two-sample t-test shows no significant difference in tissue damage score (TDS) of lesions in juxtacortical/cortical, subcortical, periventricular and brainstem regions. Both lesions meeting the inclusion criteria and confluent lesions are included in the comparisons. Each dot represents one lesion. n.s. p>0.05. All p values are after multiple comparison correction using false discovery rate.
